# Live Birth resulted from a direct thawing of vitrified human blastocyst in a regular embryo culture medium

**DOI:** 10.1101/2024.03.20.24304140

**Authors:** David Yiu Leung Chan, Murong Xu, Waner Wu, Ka Kei Fung, Crystal Hiu Yan Lock, Linyao Zhang, Hoi Man Wan, Carol Pui Shan Chan, Jacqueline PW Chung

## Abstract

**Background:** Stepwise removal of cytotoxic cryoprotectant and rehydration in serial osmotic solutions to prevent osmotic shock have been the central dogmas in cryobiology for mammalian embryos. The theory behind is to gradually remove cytotoxic cryoprotectants and rehydrate the embryo to minimize the damages during the vitrified-thaw processes. The whole process is time consuming and laborious routine in the IVF laboratory. Here, we showed that direct thawing a vitrified blastocyst in a regular human embryo culture medium without any cryoprotectant support 100% thawing survival rate regardless to blastocysts’ grading. Surprisingly, better implantation outcome was observed in our small cohort trial although not reached a significant different. There are more than million frozen embryo transfers around the world each year; a faster, safer and cheaper method can save significant amount of money for patients undergoing IVF treatment worldwide.

**Methods:** A two-phase trial were conducted: Technical phase and clinical phase. In the Technical phase, 4 commercially available one-step media and PBS solution were used for thawing procedure on donated human vitrified blastocysts. The survival rate was determined by re-expansion under time-lapse imaging. All of the one-step media at a specific range of osmolality solution between 270±10 (mOsm/KG) with human serum albumin were tested. PBS with human serum albumin at the same osmolality were used as control. We then investigated the technical comparability on routine blastocyst manipulation, we tested repeated direct thawing and repeated biopsies on the tested embryos in order to show the direct thawing method can be applied to any possible routine IVF procedures. In the clinical phase, 20 patients were recruited for the direct thawing compared to the conventional thawing of 96 patients during the same study period. Cost-effectiveness analysis was performed to estimate the different between two procedures.

**Results:** In the technical phase, we showed that all of the one-step media supported human vitrified blastocyst thawing with 100% survival rate (N=52), even in the PBS control (N=11). And the revived blastocysts are capable to re-expend comparable to conventional vitrified-thawing method. We also demonstrate this single-direct rehydrated and thawed blastocyst in embryo culture medium survived from direct thawing, trophectoderm biopsy, second vitrified, second direct thawing with the second biopsy indicated that this new method is comparable to conventional IVF procedures (N=13). Pilot clinical study showed a higher implantation rate was also obtained from this direct thawing method compared to conventional warming (61.9% VS 37.2%) and leading to healthy live birth (45% VS 36.8%). Our cost-effectiveness analysis showed that the medium direct thawing could save 42% consumable cost for patients and 90% of labor time.

**Conclusions:** We conclude that a regular embryo culture medium is comparable to the traditional thawing method that potentially can save billions of dollars and thousands of labour hours each year in IVF setting worldwide. We proved gradual cryoprotectant removal and gradual rehydration are not necessary for human blastocyst thawing survivals. Using a regular embryo culture medium for thawing blastocysts supported all clinical outcome equivalent to conventional thawing procedure. Our data also showed that the method can cut down the cost and labour time in all IVF clinics worldwide.

**Limitation:** Only single Vitrification kit and embryo carrier were used in this study. The mechanism on how the human blastocysts survived from one-step thawing remain unknown and the actual clinical power at larger scale is yet to be resolved.

## Introduction

One-third of couples worldwide experience infertility problems, and assisted reproductive technologies (ARTs) are often the only means to achieve live birth. Among all ART procedures, frozen embryo transfer (FET) is the most commonly performed assisted reproductive technology (ART) procedure since the first frozen embryo transfer by slow freezing with live birth. ^1^ Up to now, it is estimated to exceed 2 million FET cycles per year.^2^ The stepwise removal of cytotoxic cryoprotectants and rehydration in serial osmotic solutions to prevent osmotic shock has been a fundamental principle in cryobiology for decades.^3^ The theory behind this approach is to gradually eliminate cytotoxic cryoprotectants and rehydrate the embryo to minimize damage during the vitrification-thawing processes.^4-5^ The entire process is time-consuming and labor-intensive in the IVF laboratory.

Thawing of vitrified human blastocyst requires fast warming rate, a sequence of thawing solutions, and an experienced embryologist to secure the blastocyst survival rate. Currently, multiple solutions were used during blastocyst thawing from the vitrified state. The central dogma in cryobiology that several critical steps have to be fulfilled stepwise, including a high warm rate at 7700 °C/min (rapid-i, VitroLife) serial dilution of cytotoxic permeable cryoprotectant, slightly increase permeable cryoprotectant concentration for gradually rehydration of the blastocyst. It is believed that the above all factors are crucial to secure a promising survival rate in the human embryos for both cleavage and blastocyst stages. One-step thawing and rehydration without cryoprotectant in the thawing solution have never been reported feasible in the human blastocyst so far. Very recently a study led by Liebermann *et al*. showed that one-step thawing in 1M sucrose solution is possible with an encouraging higher ongoing pregnancy rate, ^6^ however; live birth has not been reported yet.

Vitrification now is the most common freezing method for blastocyst cryopreservation worldwide with the compelling survival rate compared to slow freezing. Currently, although there are several commercially available vitrification and thawing kits and numerous vitrification systems available. The basic operation and manipulation are similar among all the systems and the survival rates are comparable. Growing evidence shows that blastocyst is a very robust embryo stage that can survive from multiple biopsies and under several rounds of vitrification-thaw cycles resulted in live birth. ^7^ For a new technique or technology to be recognized by in field professionals, there are two main issues have to be investigated beforehand. They are the efficiency and safety. Vitrification-thawing method has been demonstrated a better method over slow freezing in terms of embryo survival, implantation, pregnancy (in both biochemical and clinical) and live birth rate. Up to now, long-term safety is yet to be proven but preliminary outcomes on live birth data did not show any serious adverse events in terms of prenatal or postnatal measurements.

Here, we demonstrate, for the first time, a direct thawing and rehydration using a regular embryo culture medium, without any intermediate solution with cryoprotectant that supports 100% thawing survival rate in a single step regardless of blastocyst grading. Surprisingly, our cohort trial showed improved live birth outcomes. This faster, safer, and more cost-effective procedure has the potential to save billions of dollars each year for patients undergoing IVF treatment worldwide.

## Materials and methods

### IRB approval

The research report described herein has been carried out in accordance with The Code of Practice of the Hong Kong Human Reproductive Technology Council and the Code of Ethics of the World Medical Association (Declaration of Helsinki) for studies involving humans. This study was approved by the Hospital research board of the Faculty of Medicine, Prince of Wales Hospital, The Chinese University of Hong Kong (CREC Ref. No. 2010.432 and IRB/REC Ref. No. 2023.066). Written informed consent was obtained from the patient for the study and publication.

### Donated blastocysts for the technical phase research

All blastocysts used in this study had previously been donated and stored for research purposes at the ART unit in department of obstetrics and gynaecology, The Chinese University of Hong Kong. All the patients information were remained anonymous.

### Blastocyst vitrification

All the donated blastocysts were originally from surplus embryos and later decided to be discarded. Blastocysts were laser shrunk and then vitrified using Irvine vitrification kit (Product no.90188 or previous version 90133-OS) on Rapid-i (VitroLife product no. 14406) vitrification straw for store until experiments were performed. In brief, blastocysts were incubated in 50ul ES solution for ten minutes and then to VS solution for less than 90 seconds. The blastocyst was then loaded on the straw and plunged into the sleeve which was pre-cool in liquid nitrogen for at least two minutes prior to the straw insertion. The sleeve was then sealed and store in LN tank for long term storage until use.

### Embryo culture condition, time lapse imaging and blastocyst grading

Gardner’s grading system were used on accessing blastocyst status based on morphology. All of the embryos were cultured in VitroLife G-IVF plus medium (Product no. 10134) on the day of oocyte pick up (OPU) and insemination. After fertilization checking normal fertilized zygotes were then transferred into VitroLife G-TL plus medium (Product no. 10145) covered with Ovoil (product no. 10029) until day 5 or 6. All embryo culture were incubated in either VitroLife EmrbyoScope or Cook Minc incubator at 37oC with 6% of CO2, 5% O2 and 6% of CO2 and atmospheric pressure respectively.

### Single step thawing in medium

Vitrified blastocysts were thawed directly into 1ml G-TL medium (VitroLife, product no. 10145), Single step culture medium (Kitazato, SK-FCBS-50), Continuous single culture-NX complele medium (Fujifilm-Irvine, 90168) and Globle Total medium (Life Global, LGGT-030) at 37oC in a Nunc organ dish (Corning 353037). Rapid-i embryo carrier was removed from LN tank and the sleeve was cut open. The embryo containing straw were taken out from the sleeve and directly submerged into to 1 ml medium/tested solution which were pre-incubated in Cook incubator in 6% CO2 at 37°C overnight (for carbonate base medium only) or pre-warm to 37 °C before use. After 1 minute, the blastocyst was transfer to fresh VitroLife G-TL medium and incubate in time-lapse incubator for subsequent observation and experiments. The thawing video is available in the supplementary video 1.

### Conventional thawing using thawing kit

Vitrification thawing kits were obtained from Irvine Scientific (90137-SO) and followed the user instructions. The embryo containing Rapid-i sleeve was cut opened and the straw was taken out and dipped in 1 ml TS solution at 37°C, immediately transferred to a 50 ul fresh TS solution for 2 minutes, DS solution twice for 4 minutes and WS solution three times for total 9 minutes. The whole conventional thawing method takes approximately 18-20 minutes.

### Direct-thaw, trophectoderm biopsy, re-vitrify, re-thaw and re-biopsy

Trophectoderm biopsies and tubing of the tissue were performed by experienced embryologists in our centre. RI Integra 3 manipulator and Saturn 5 laser system were used for the procedures. 5-10 cells of the trophectoderm were taken from the blastocysts, biopsy needle (Cooper surgical, MBB-FP-SM-30) and holding pipette (Cooper surgical, MPH-MED-30) were used. The biopsied blastocysts were then re-vitrified under conventional method and then a second time thawing was performed. Re-thawed blastocysts were observed under time lapse system until expended and then second biopsies were obtained.

### The pilot direct thawed frozen embryo transfer and clinical outcome

20 Patients pending for FET were consented via hospital consent form satisfying the Hong Kong council for human reproductive technologies and local regulations (Cap. 561). The blastocysts were thawed as above mentioned in the VitroLife G-TL medium for embryo transfer. Cook Guardia embryo transfer catheters (K-JETS-7019-ET) were used for the embryo transfer. Approximately 2-5 ul medium with the embryo, medium column surrounded by two air gaps on each end and then the embryo was ejected gently into the uterine cavity. A blood sample was taken 9 days after transfer of a blastocyst for human chorionic gonadotropin (HCG) measurement to verify if pregnancy had occurred. In women with a positive pregnancy test, a transvaginal ultrasound scan was performed 2 weeks later to determine the location of pregnancy, the number of gestational sacs and fetal viability. The fetal heart beat was defined as a clinical pregnancy. All of the cases were follow-up as standard practice under our hospital clinical protocol.

### Statistical analysis

Comparison between categorical variables expressed in percentages (%) was performed with student t-test for independent samples. Statistical significance was set at P < 0.05. Statistical analysis was performed with the SPSS Statistics programme (version 27, IBM Corp., New York, USA).

## Results

### Technical Phase

In this phase, we tested 76 blastocysts on direct thawing in 5 different media and showed the compatibility on repeated direct thawing and biopsies. Figure 1 showed the schematic flow of this study.

**Figure 1.**
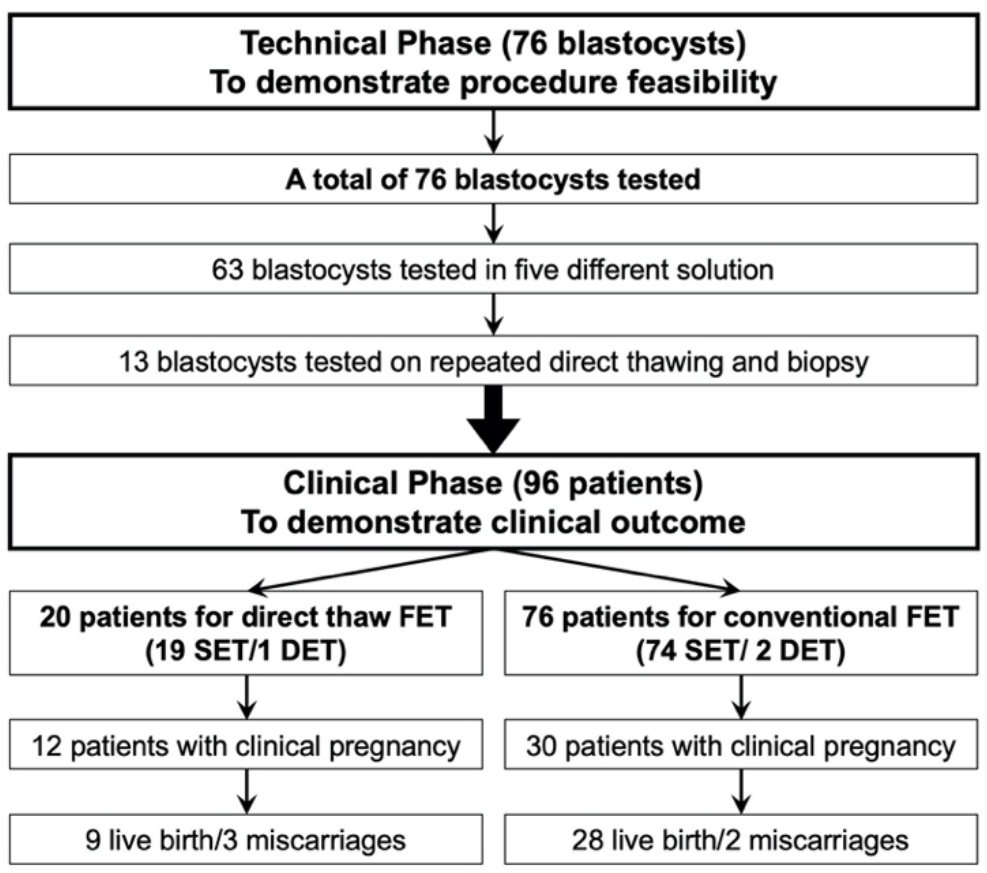
Schematic flowchart of the study

### 100% blastocysts survival rate under direct thawed in all tested media

We tested 63 donated human blastocysts in total 5 different media (Figure 2b). We demonstrated 100% survival rate when blastocysts thawed in all the media without stepwise procedures. This finding challenges the central dogma of cryobiology on human blastocyst. The vitrified blastocyst-containing carrier was immersed in a pre-warmed medium at 37 °C and incubated for one minute at a heat-top workstation before being transferred to a time-lapse incubator for further observation. Time-lapse imaging showed that all blastocysts survived the direct thawing procedure and were able to re-expand (Figure 2a and suppl. Video 1). The re-expansion of the blastocoels, without the use of any invasive markers, indicated their survival. Additionally, we tested all the commercially available one-step embryo culture media alongside a Phosphate buffered saline (PBS) only control (N=11). Interestingly, all blastocysts survived at an osmolality of 270±10 mOsm/kg in the PBS solution.

**Figure 2.**
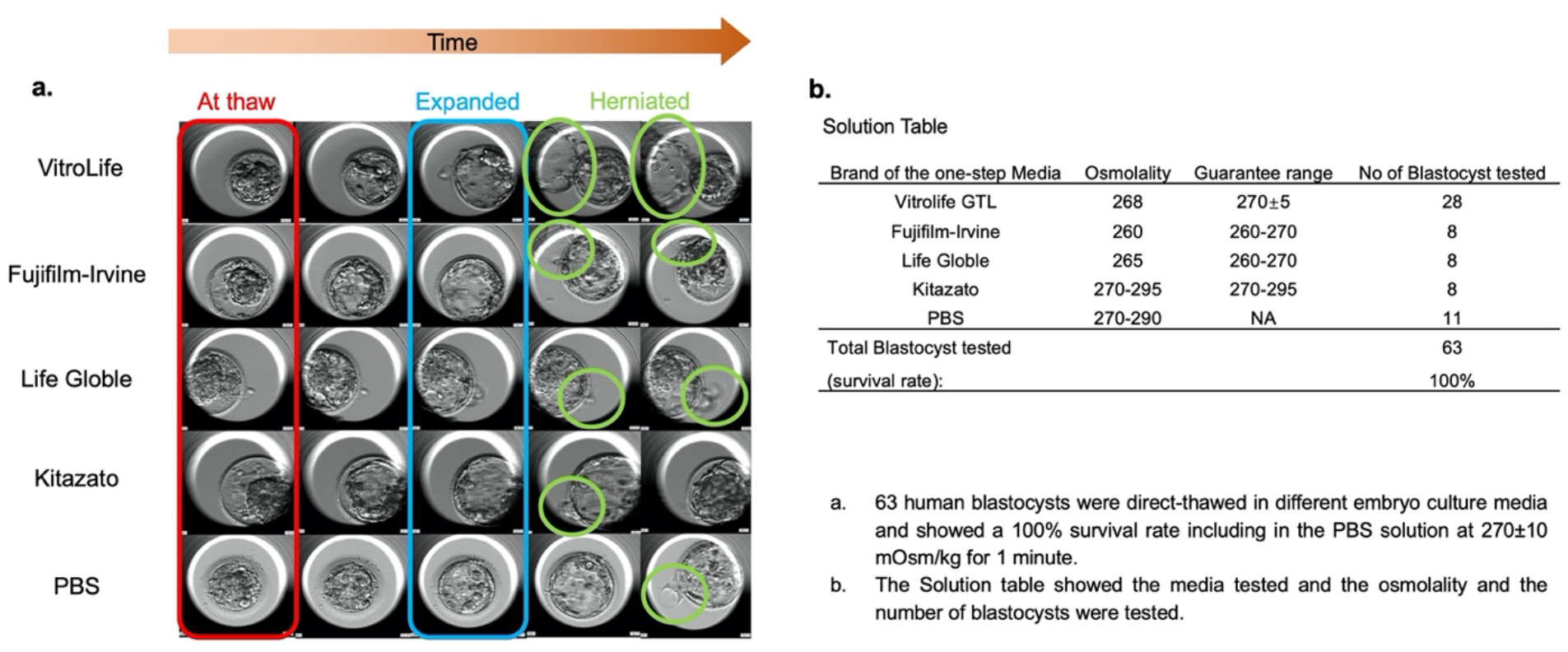
Direct thawed embryo showed normal re-expansion and herniate features

### Direct thawing in medium can survive repeated thawing and biopsies

Conventional ART treatments often involve multiple vitrify-thaw cycles, such as cleavage stage frozen-thaw to day 5-6 blastocyst or for preimplantation genetic testing (PGT) biopsies. We tested 13 donated blastocysts for vitrified-direct-thaw biopsies and re-vitrified, re-direct-thaw re-biopsy procedures. Figure 3a showed the schematic diagram on the repeated procedures. All blastocysts survived in double direct thawing, and double biopsies showed that the direct thawing method is comparable to conventional multiple vitrified-thawed procedures in modern ART laboratories (Figure 3b). In summary, the technical phase demonstrated high survival and compatibility on IVF procedures.

**Figure 3.**
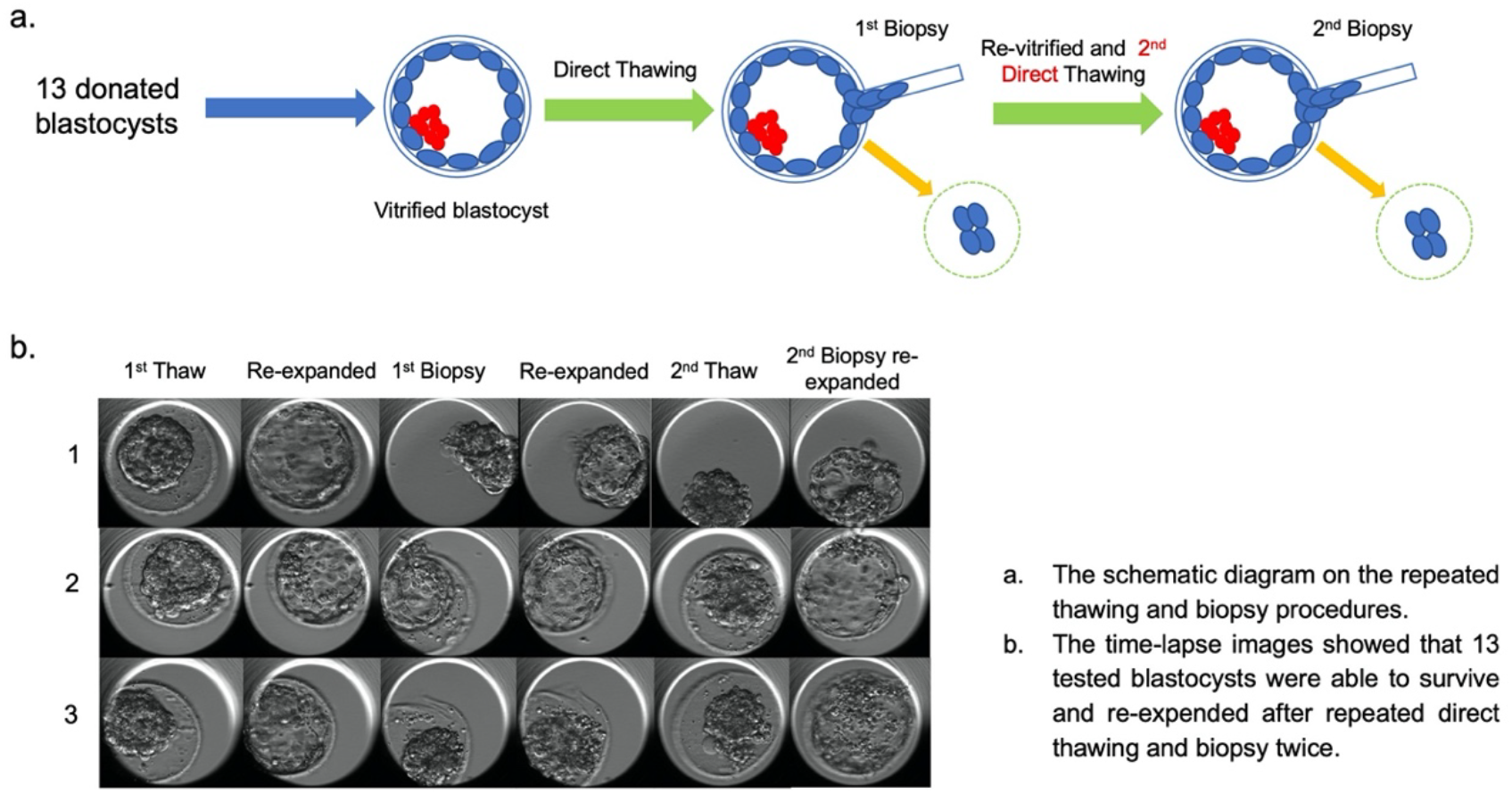
Schematic diagram on repeated thawing and biopsy

### Clinical Phase

#### Direct thawing in medium supported pregnancy and live birth

We then conducted a proof-of-concept trial on the direct thawing method, involving a cohort of 96 patients who were scheduled for frozen embryo transfers. Patients’ demographics are showed in the table 1. 20 patients were recruited for direct thawing FET. Among 20 patients under direct thaw FET, 19 were single embryo transfer (SET) while one was double embryo transfer (DET). In the conventional thawing group with 76 patients, 74 with SET and 2 with DET. All of the thawed blasocysts were survived from both of the thawing procedures. The implantation rate in the direct thawing group was significant higher at 61.9% (13 sacs/21 blastocysts) compared to the conventional method at 37.2% (29/78). Although the live birth rate was also higher in the direct thawing group at 45% (9 LB/20 ETs) compared to 36.8% (28 LB/78 ETs) in the conventional thawing group, the difference was not statistically significant (Figure 4a). All nine live births were healthy without any neonatal complications or abnormalities. This pilot trial demonstrated that the direct thawing procedure in embryo culture medium is superior to the conventional method in terms of pregnancy outcomes, and live birth rate. The miscarriage rate was also non-significant between two groups.

**Table 1.** Patient demographic data:

|  | Value | P Value |
| --- | --- | --- |
| Age at the OPU date | 35.39±0.74 | .147<sup>a</sup> |
| Number of embryos observed | 1.65±0.19 | .000<sup>a</sup> |
| Diagnosis for infertility |  |  |
| Primary | 51 (58%) |  |
| Secondary | 37 (42%) |  |
| Male Indication |  |  |
| No male indication | 36 (40.9%) |  |
| Borderline Male Factor | 6 (6.8%) |  |
| Moderate/Severe Male Factor | 45 (51.1%) |  |
| Coital Dysfunction | 1 (1.1%) |  |
| Stimulation protocol |  |  |
| Antagonist (Flexible) | 39 (44.3%) |  |
| Antagonist pre-treatment OC | 44 (50%) |  |
| Long Protocol/ Down Regulated | 5 (5.7%) |  |
| Number of follicles | 14.31±0.96 | .189<sup>a</sup> |
| Number of OCC | 13.13±1.18 |  |
| Number of fertilized oocytes | 8.92±0.86 |  |
| Number of cleaved embryos | 8.78±0.84 |  |
| Fertilization rate | 69.0% |  |
| Blastocyst formation rate | 66.0% |  |
| Pregnancy rate | 46.8% |  |
| Cumulative Pregnancy rate | 68.8% |  |
| Live Birth rate | 31.9% |  |
| Cumulative Live Birth rate | 51.3% |  |

a Shapiro-Wilk Test for the Normal distribution
b Median for data that is not normally distributed

**Figure 4.**
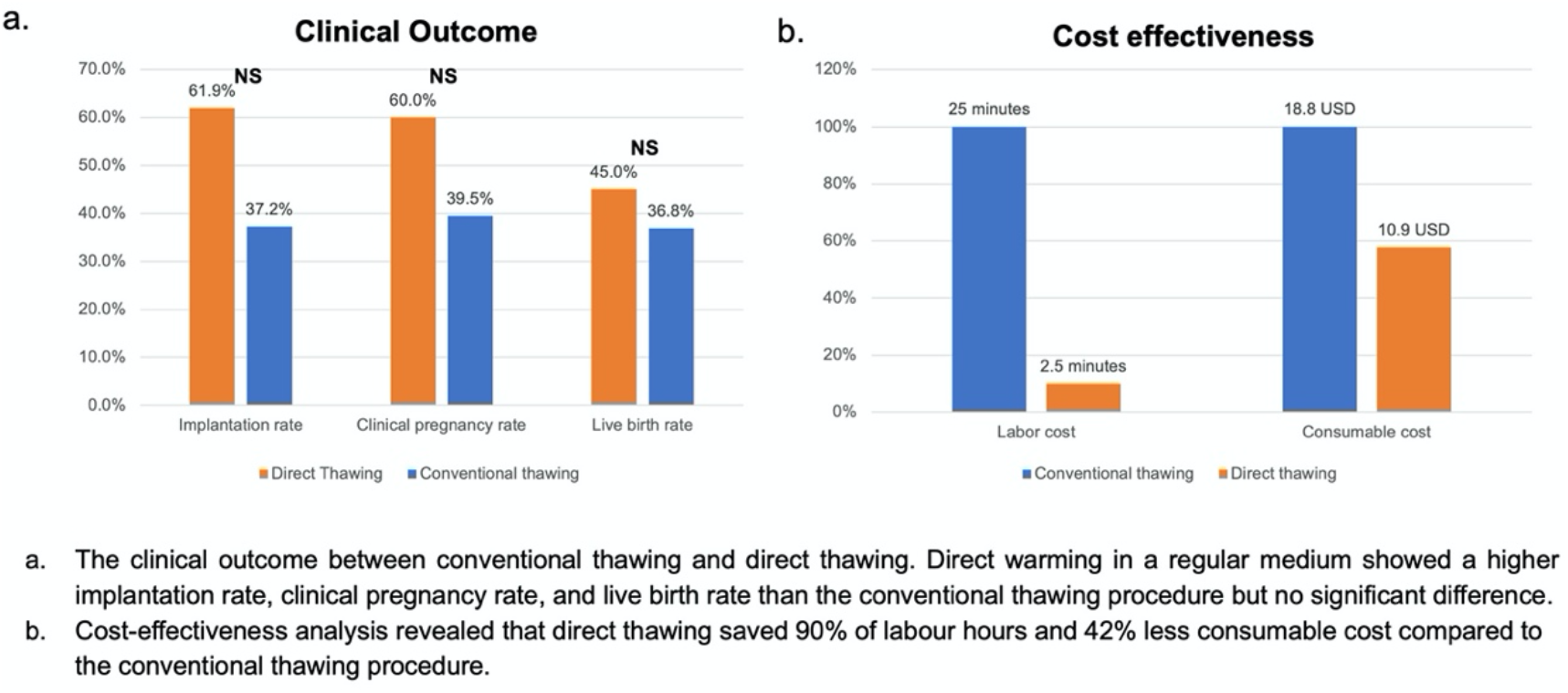
Clinical outcome and cost effectiveness analysis

#### Cost-effectiveness analysis

The direct thawing procedure in embryo culture medium obviously can reduce lab cost and labour hour per frozen embryo transfer. We conducted cost-effectiveness analysis and estimated a significant reduction in labor costs of up to 90% and a 42% reduction in consumable costs per frozen embryo transfer (Figure 4b).

## Discussion

Multiple steps for blastocyst thawing is a regular procedure in ART laboratory, it involves several steps first to remove of cytotoxic cryoprotectant and gradually re-hydrate the blastocyst from the vitrified state. It is believed that the multi-step thawing procedure is a must to secure the survival rate, any deviation from the procedure will jeopardize the rapid reviving blastocyst. Thawing of blastocysts by conventional method required an experienced embryologist to manipulate the blastocyst in multiple solutions during the thawing. The thwaing is time consuming and labor intensive. A single-step thawing method without pipetting embryos from drop to drop can minimize the risk of losing the blastocyst during manipulation and miminize the time in suboptimal culture condition. Furthermore, single-step thawing procedure significantly shorten the time of thawing and less consumable used. A recent study led by Liebermann *et al* demonstrated a single step thawing in 1.0M sucrose solution with higher ongoing pregnancy rate. ^6^ The research is a big leap for achieving one step thawing on human embryos but live birth data is not available yet. Our data suggested that a simple single-step thawing procedure in a regular embryo culture medium, without intermediate cryoprotectant with excellent survival rate leading to live births. Our data also showed that blastocysts survived in all of the tested commercial media at the specific range of osmolality from 270±10 including the PBS solution. Implying the essential human embryo culture medium provides enough support for the direct thawing and rehydration. The direct thawing incubation time is also an important factor for the blastocyst to survive; although we did not check the optimum time for the direct thawing procedure due to limited number of donated blastocysts; it seemed 1 minute was the best for all of the tested media.

Vitrification has been used in ART laboratory for more than two decades. Many groups tried to design a faster and less technically challenging procedure for embryologists while aimed to retain equivalent or better embryo outcome. Direct thawing for animal embryos are using for decades, however; has never been used in human embryos given the ethical issue and limited donated materials for further research. Previous one-step dilution was reported successfully in bovine blastocyst; ^7^ however the thawing was not purely single-step involving thawing from room temperature for 8-15 seconds and then in 37°C pre-warm water bath; together with increased serum level in the medium. Also, the presence of opening on the zona pellucida contributed to the survival. Compared to our study, although in much smaller sample size due to the ethical and recruitment issues; we showed that a single dip in a solution at 270±10 mOsm/kg in a second, either with or without opening on the zona pellucida, resulted in 100% survival rate. It has been demonstrated that abnormal fertilized human oocytes with zero survival upon direct rehydration in culture medium at 38°C, ^8^ indicated that hypo-osmotic stress was detrimental to one cell embryo given the large cell size.

RCT is needed to show the pregnancy outcome in a larger population. The limitation of this study is the long term health effects after live birth is also not yet available. Long time follow up to reassure long term technical safety from this new method.

## Conclusion

Taken together, we conclude that the direct thawing in an ordinary embryo culture medium is comparable to the conventional-serial thawing method. The direct thawing procedure has the potential to increase pregnancy rates and live birth rates while saving billions of dollars and thousands of labor hours each year in IVF settings worldwide, especially in low resources area. We have demonstrated that gradual cryoprotectant removal and gradual rehydration are not necessary for blastocyst thawing survival if the osmolality is maintained within the specific range. The one-step, direct thawing procedure in an ordinary embryo culture medium can significantly reduce costs and labor time in all IVF clinics worldwide.

## Data Availability

Our data are available with a signed data use agreement; please contact.

## Acknowledgments

We would like to thank the Hong Kong Health and Medical Fund (Ref 18190321), from the Hong Kong government, The Hong Kong Research Matching Grant (RMG01-8601386) to support this clinical research. A Special gratitude to my IVF team members to support innovative research eventually could reduce their burdens in the near future.

## Appendix

Supplemental Video 1: Direct thawing method illustration and culturing time-lapse video (self-explanatory);

## Data availability

Our data are available with a signed data use agreement; please contact.

## References

1. Trounson A, Mohr L: Human pregnancy following cryopreservation, thawing and transfer of an eight-cell embryo. Nature 1983, 305:707–9.

2. Adamson GD Z.-H.F., Dyer S, Chambers G, de Mouzon J, Ishihara O, Kupka M, Banker M, Jwa SC, Elgindy E, Baker V.. ICMART preliminary world report 2018. 2022; Available from: https://www.icmartivf.org/wp-content/uploads/ICMART-ESHRE-WR2018-Preliminary-Report.pdf

3. Rall WF, Fahy GM: Ice-free cryopreservation of mouse embryos at -196 degrees C by vitrification. Nature 1985, 313:573–5.

4. Kader AA, Choi A, Orief Y, Agarwal A: Factors affecting the outcome of human blastocyst vitrification. Reprod Biol Endocrinol 2009, 7:99.

5. Wilmut I: The effect of cooling rate, warming rate, cryoprotective agent and stage of development on survival of mouse embryos during freezing and thawing. Life Sci II 1972, 11:1071–9

6. J. Liebermann, K. Hrvojevic, J. Hirshfeld-Cytron, R. Brohammer, Y. Wagner, A. Susralski, et al. Fast and furious: pregnancy outcome with one-step rehydration in the warming protocol for human blastocysts; Reproductive BioMedicine Online 2024 Vol. 48 Issue 4 Pages 103731

7. S. Zhang, K. Tan, F. Gong, Y. Gu, Y. Tan, C. Lu, et al.; Blastocysts can be rebiopsied for preimplantation genetic diagnosis and screening; fertil Steril 2014 Vol. 102 Issue 6 Pages 1641–5

8. V. Isachenko, M. Montag, E. Isachenko, F. Nawroth, S. Dessole and H. van der Ven Developmental rate and ultrastructure of vitrified human pronuclear oocytes after step-wise versus direct rehydration; human Reproduction 2004 Vol. 19 Issue 3 Pages 660–665

9. G. Vajta, C. N. Murphy, Z. Macháty, R. S. Prather, T. Greve and H. Callesen In-straw dilution of bovine blastocysts after vitrification with the open-pulled straw method; Vet Rec 1999 Vol. 144 Issue 7 Pages 180–1 Accession Number: 10097328

